# Antibiotic De-escalation Patterns and Outcomes in Critically Ill Patients with Suspected Pneumonia as Informed by Bronchoalveolar Lavage Results

**DOI:** 10.1101/2024.09.10.24313149

**Authors:** Mengou Zhu, Chiagozie I. Pickens, Nikolay S. Markov, Anna Pawlowski, Mengjia Kang, Luke V. Rasmussen, James M. Walter, Nandita R. Nadig, Benjamin D. Singer, Richard G. Wunderink, Catherine A. Gao, The NU SCRIPT Study Investigators

**Affiliations:** Department of Medicine, Northwestern University Feinberg School of Medicine, Chicago, IL; Division of Pulmonary and Critical Care, Northwestern University Feinberg School of Medicine, Chicago, IL; Northwestern Medicine Enterprise Data Warehouse, Chicago, IL; Division of Health and Biomedical Informatics, Northwestern University Feinberg School of Medicine, Chicago, IL

## Abstract

**Background:** Antibiotic stewardship in critically ill pneumonia patients is crucial yet challenging, partly due to the limited diagnostic yield of noninvasive infectious tests. In this study, we report an antibiotic prescription pattern informed by bronchoalveolar lavage (BAL) results, where clinicians de-escalate antibiotics based on the combination of quantitative cultures and multiplex PCR rapid diagnostic tests.

**Methods:** We analyzed data from SCRIPT, a single-center prospective cohort study of mechanically ventilated patients who underwent a BAL for suspected pneumonia. We used the novel Narrow Antibiotic Therapy (NAT) score to quantify day-by-day antibiotic prescription pattern for each suspected pneumonia episode etiology (bacterial, viral, mixed bacterial/viral, microbiology-negative, and non-pneumonia control). We also analyzed and compared clinical outcomes for each pneumonia etiology, including unfavorable outcomes (a composite of in-hospital mortality, discharge to hospice, or requiring lung transplantation during hospitalization), duration of ICU stay, and duration of intubation. Clinical outcomes were compared with the Mann-Whitney U test and Fisher’s exact test.

**Results:** We included 686 patients with 927 pneumonia episodes. NAT score analysis indicated that an antibiotic de-escalation pattern was evident in all pneumonia etiologies except resistant bacterial pneumonia. Microbiology-negative pneumonia was treated similarly to susceptible bacterial pneumonia in terms of antibiotic spectrum. Over a quarter of the time in viral pneumonia episodes, antibiotics were completely discontinued. Unfavorable outcomes were comparable across all pneumonia etiologies. Patients with viral and mixed bacterial/viral pneumonia had longer durations of ICU stay and intubation.

**Conclusions:** BAL quantitative cultures and multiplex PCR rapid diagnostic tests resulted in prompt antibiotic de-escalation in critically ill pneumonia patients. There was no evidence of increased incidence of unfavorable outcomes.

**Key points:** We found that bronchoalveolar lavage-based diagnostics is associated with antibiotic de-escalation, including to no antibiotics in more than a quarter of cases where only a virus is recovered.

## Introduction

Pneumonia is a leading cause of morbidity and mortality worldwide.[1] Antibiotic stewardship and de-escalation for pneumonia in critically ill patients is crucial given the rise of multi-drug resistant (MDR) organisms.[2] Despite the most recent guidelines from the Infectious Diseases Society of America (IDSA) advocating for antibiotic de-escalation in patients with no evidence of resistant organisms, substantial provider hesitancy in antibiotic de-escalation exists, even when the microbiology workup yields negative results.[3] Such provider hesitancy is partly due to the limited diagnostic yield of infectious workup for bacterial pneumonia, as well as concerns for patient safety and outcomes.

Current IDSA guidelines recommend noninvasive respiratory tract sampling, such as sputum sample and endotracheal aspirate, to identify causative organisms in patients with community-acquired pneumonia (CAP) who are at risk for resistant organisms or severe disease, as well as in patients with hospital-acquired pneumonia (HAP) and ventilator-associated pneumonia (VAP).[4,5] However, high-quality, uncontaminated sputum samples can only be successfully obtained from a limited percentage of patients with CAP and HAP.[4,6,7] In a study of hospitalized patients with CAP, only 62% could provide a sputum sample, out of which only 57% met criteria for high-quality specimens.[6] In a study of patients with HAP, only 29.4% of patients had a sputum sample collected, out of which 64% were contaminated.[7] Sputum Gram stains have a rapid turnaround time, but the sensitivity is suboptimal. For example, a positive stain for Gram-positive diplococci is only 68.2% sensitive for identifying *S. pneumoniae.*[6] For patients with VAP, endotracheal aspirate can sometimes show discordant Gram stain and semiquantitative culture results with lower respiratory tract samples, indicating limited diagnostic accuracy. In a study focusing on the concordance between diagnostic workup performed on endotracheal aspirate and bronchoalveolar lavage (BAL) fluid, Gram stain results on endotracheal aspirates and BAL fluid only agreed about 60% of the time, and semiquantitative cultures performed on endotracheal aspirates can have a sensitivity as low as 38%.[8] Blood cultures are another commonly used method of noninvasive infectious sampling, but their diagnostic yield is low in pneumonia regardless of the severity and presence of risk factors for resistant organisms.[4,9] Invasive sampling of the lower respiratory tract with BAL often has better diagnostic yields, but such tests are performed at variable rates (proportions ranging from 0% to 25.8% of mechanically ventilated patients in one study),[10] even in critically ill patients.[11–13] BAL samples can be the substrate of multiple tests crucial to diagnosing and characterizing pneumonia, including cell count and differential, quantitative cultures, and multiplex PCR rapid diagnostic tests (RDT). While not included in current pneumonia guidelines, multiplex PCR RDTs performed on BAL fluid provide rapid results and effectively identify infectious organisms.[14–16]

Ideally, antibiotic de-escalation in critically ill patients with pneumonia should be based on a rapid, sensitive, and specific microbiology diagnostic test. Combining multiplex PCR RDTs and quantitative cultures performed on BAL samples can potentially achieve such diagnostic goals and can be incorporated into antibiotic de-escalation strategies.[17] When given this sensitive and detailed data, clinicians in our center often de-escalate antibiotics.[11] Various ways of quantifying antibiotic administration have been proposed such as days of therapy.[18] Here we use the Narrow Antibiotic Therapy (NAT) score, which quantifies the breadth of antibiotic coverage as well as days administered.[11] In this study, we describe the antibiotic prescription pattern across different types of pneumonia (i.e., bacterial, viral, mixed bacterial/viral, microbiology-negative) in critically ill pneumonia patients when antibiotic de-escalation is informed by the combination of multiplex PCR RDT and quantitative cultures performed on BAL samples.

## Methods

### Study Cohort

Patients were enrolled in the Successful Clinical Response In Pneumonia Therapy (SCRIPT) study (IRB STU00204868), a single-center prospective cohort study of patients with suspected pneumonia admitted to the medical intensive care unit (MICU) of Northwestern Memorial Hospital and requiring mechanical ventilation.[11,19,20] Patient families or legally authorized representatives consented to participate in the study. If a patient was readmitted to the hospital, they were treated as a new study subject and given a new identifier. The SCRIPT research team reviewed patient charts on study enrollment to determine who fulfills set criteria of immunocompromised conditions (see Table 1 footnote).

**Table 1.** Demographics and Clinical Characteristics in All Patients.

|  | **Overall (N=686)** |
| --- | --- |
| **Age** |  |
| **Median [Q1, Q3]** | 62.0 [51.0, 71.0] |
| **Gender (male)** | 409 (59.6%) |
| **Race** |  |
| **Asian** | 22 (3.2%) |
| **Black or African American** | 134 (19.5%) |
| **Unknown or Not Reported** | 125 (18.2%) |
| **White** | 405 (59.0%) |
| **Ethnicity** |  |
| **Hispanic or Latino** | 140 (20.4%) |
| **Not Hispanic or Latino** | 514 (74.9%) |
| **Unknown or Not Reported** | 32 (4.7%) |
| **BMI** |  |
| **Median [Q1, Q3]** | 28.5 [24.2, 33.7] |
| **Missing** | 6 (0.9%) |
| **Immunocompromised\*** | 206 (30.0%) |

### Pneumonia Episodes

BAL was performed for suspected pneumonia episodes at the discretion of the treating MICU team. At our center, BAL samples were routinely acquired to guide clinical decision-making. BAL is either performed by physicians through fiberoptic bronchoscopy or by respiratory therapists through a non-bronchoscopic approach (NBBAL). BAL samples were sent for routine clinical tests including cell count and differential, quantitative culture, and many were sent for multiplex PCR RDT (BioFire^®^ FilmArray^®^ Pneumonia Panel) to identify possible causative organisms. A panel of critical care physicians reviewed all available clinical data from the electronic health record (EHR) and adjudicated suspected pneumonia episodes based on a pre-determined multi-reviewer protocol.[21] They categorized each episode into bacterial pneumonia, viral pneumonia, mixed bacterial/viral pneumonia, microbiology-negative pneumonia, and non-pneumonia control based on the BAL results and chart review.

Microbiology-negative pneumonia was adjudicated based on pulmonary-specific features such as chest imaging, BAL neutrophilia, and ventilator parameters, and required clinical consensus from multiple reviewers. Presence of extra-pulmonary infection was also reviewed based on each patient’s EHR. Each bacterial pneumonia episode was further adjudicated on day 7 following BAL to determine whether the pneumonia had been clinically cured based on improvements in features such as fever curve, leukocytosis, and ventilator settings. Specific details of the adjudication protocol and criteria for clinical cure have been published.[21] Bacterial pneumonia episodes were categorized into resistant and susceptible based on the results of quantitative culture and multiplex PCR. An episode was defined as resistant if the causative organism tested positive for any antibiotic resistance gene (*mec*A/C, MREJ, OXA-48-like, CTX-M, KPC, NDM, IMP, VIM), or if the quantitative culture result indicated a resistant pathogen (such as an ESBL-positive organism or MRSA). The code for processing PCR and quantitative culture results can be found at https://github.com/NUSCRIPT/mz_abx_deescalation_2024.

### Characterizing Antibiotic Stewardship Pattern with the Narrow Antibiotic Treatment (NAT) Score

We quantified the spectrum of antibiotic therapy administered to each patient on each day using the NAT score. Briefly, a score of 0 corresponded to empiric guideline-recommended CAP coverage of ceftriaxone and azithromycin, -1 corresponded to monotherapy with a narrow-spectrum antibiotic, and -2 corresponded to no antibiotic treatment. Antibiotic therapies with broader spectrums, such as for resistant HAP or VAP, were assigned higher scores.[11,22] A description and examples of how NAT scores were assigned and analyzed relative to post-BAL days are included in Figure 1.

**Figure 1.**
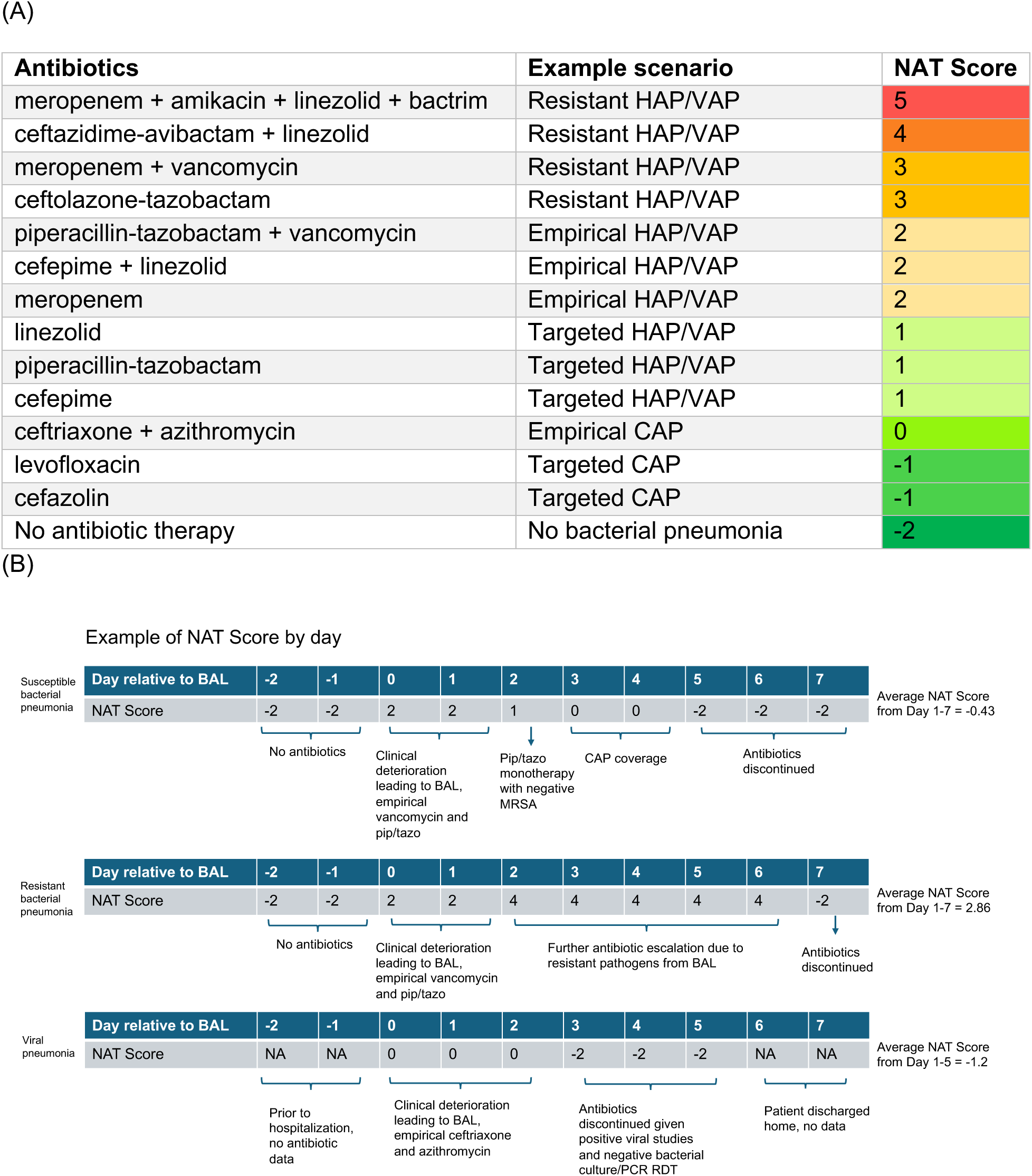
(A) Examples of the NAT score scenarios and (B) day by day examples of different clinical scenarios. The example NAT table and scenarios are adapted from our group’s prior work, references [11] and [22].

Antibiotic therapy spectrum for each pneumonia etiology was visualized by plotting the median and interquartile range (IQR) of NAT score against day, from two days before to seven days after the episode-defining BAL. If a patient died or was discharged less than seven days after the episode-defining BAL, then only the NAT scores during the inpatient days were included. Antibiotic de-escalation patterns were visualized by plotting the average NAT score over the first to the seventh day after the episode-defining BAL, or to the day of death or discharge, whichever came first.

### Patient Outcomes

Clinical outcomes in patients with pneumonia due to different etiologies were analyzed and compared. For outcome analysis, we only included the patients with exactly one suspected pneumonia episode in order to avoid concurrent contribution from multiple different pneumonia etiologies to patient outcomes. We studied unfavorable outcomes (a composite of in-hospital mortality, discharge to hospice, or requiring lung transplantation during hospitalization), duration of ICU stay, and duration of mechanical ventilation. The patient cohort flow diagram is presented in Figure 2.

**Figure 2.**
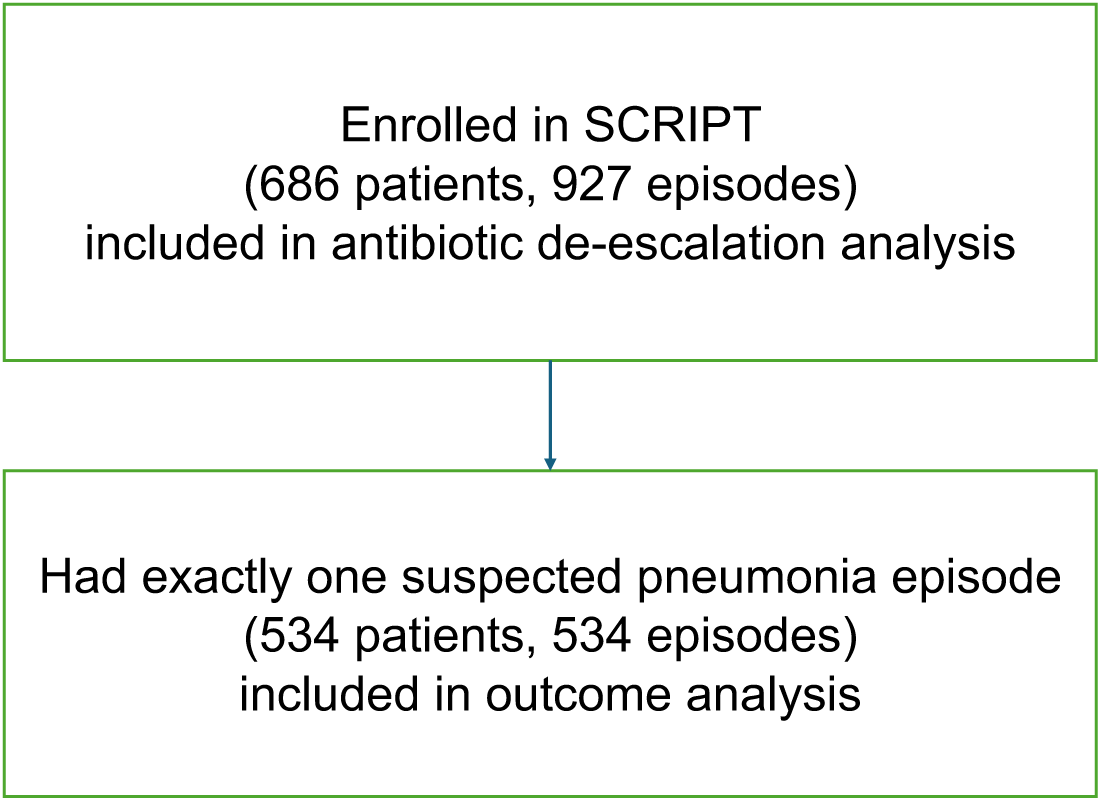
Flow diagram of patients included in the different analyses. All patients with suspected pneumonia episodes were included in the antibiotic de-escalation analysis. Patients who had exactly one suspected pneumonia episode was included in the outcomes analysis.

### Statistical Analysis

Average NAT scores were compared using Mann-Whitney U tests, with p<0.05 as the cutoff for statistical significance. Patient mortality across different pneumonia etiologies was compared using Fisher’s exact test. Duration of ICU stay and duration of mechanical ventilation were compared using Mann-Whitney U tests. Statistical analyses and plotting were performed with R version 4.3.1. Data were analyzed with dplyr version 1.1.2 and tidyr version 1.3.0. All plots were generated with ggplot2 version 3.4.2 and ggpubr 0.6.0. Preprocessing was performed with Python version 3.9. Code is available at repository https://github.com/NUSCRIPT/mz_abx_deescalation_2024.

## Results

### Clinical Characteristics of Study Cohort

686 patients enrolled in SCRIPT between June 2018 and April 2023 had complete clinical adjudication information at the time of analysis (Table 1). Among these patients, 409 (59.6%) were male, the median age was 62 years, and 206 (30.0%) were immunocompromised. These patients had 927 clinical episodes that were subsequently adjudicated to be bacterial pneumonia (n = 288), viral pneumonia (n = 176), mixed bacterial/viral pneumonia (n = 198), microbiology-negative pneumonia (n = 151), and non-pneumonia control (n = 114). These episodes were included in the visualization and analysis of antibiotic stewardship patterns. Of these episodes, 150 episodes were adjudicated as CAP, 257 as HAP, and 406 as VAP. Of the viral and mixed bacterial/viral pneumonia cases, 76% cases were SARS-CoV-2. Of the 486 bacterial and bacterial/viral episodes, 104 had resistant pathogens as identified using our logic (Supplemental Table 1). The organisms detected are presented in Supplemental Table 2. A detailed breakdown of all pneumonia episodes by category and etiology is summarized in Supplemental Table 3. The frequencies of other viruses causing pneumonia episodes can be found in Supplemental Table 4.

Of all 686 patients, 534 had exactly one suspected pneumonia episode and were included in the clinical outcome analysis. Out of these patients, 164 had bacterial pneumonia, 93 had viral pneumonia, 76 had mixed bacterial/viral pneumonia, 107 had microbiology-negative pneumonia, and 94 were adjudicated not to be pneumonia (non-pneumonia control) (Table 2).

**Table 2.** Demographics and Clinical Characteristics in Patients with Exactly One Suspected Pneumonia Episode, Separated by Pneumonia Category.

|  | Bacterial | Viral | Bacterial/viral | Microbiology-negative | Non-pneumonia Control | Overall |
| --- | --- | --- | --- | --- | --- | --- |
|  | (N=164) | (N=93) | (N=76) | (N=107) | (N=94) | (N=534) |
| **Age (Median [Q1, Q3])** | 66.0 [50.8, 73.3] | 61.0 [50.0, 69.0] | 62.5 [52.0, 71.3] | 61.0 [51.5, 73.0] | 58.5 [49.3, 69.8] | 62.0 [51.0, 72.0] |
| **Gender (male)** | 108 (65.9%) | 50 (53.8%) | 48 (63.2%) | 53 (49.5%) | 46 (48.9%) | 305 (57.1%) |
| **Race** |  |  |  |  |  |  |
| **Asian** | 4 (2.4%) | 2 (2.2%) | 3 (3.9%) | 1 (0.9%) | 1 (1.1%) | 11 (2.1%) |
| **Black or African American** | 38 (23.2%) | 15 (16.1%) | 13 (17.1%) | 23 (21.5%) | 17 (18.1%) | 106 (19.9%) |
| **Unknown or Not Reported** | 24 (14.6%) | 25 (26.9%) | 14 (18.4%) | 14 (13.1%) | 14 (14.9%) | 91 (17.0%) |
| **White** | 98 (59.8%) | 51 (54.8%) | 46 (60.5%) | 69 (64.5%) | 62 (66.0%) | 326 (61.0%) |
| **Ethnicity** |  |  |  |  |  |  |
| **Hispanic or Latino** | 15 (9.1%) | 32 (34.4%) | 28 (36.8%) | 12 (11.2%) | 13 (13.8%) | 100 (18.7%) |
| **Not Hispanic or Latino** | 143 (87.2%) | 57 (61.3%) | 42 (55.3%) | 89 (83.2%) | 77 (81.9%) | 408 (76.4%) |
| **Unknown or Not Reported** | 6 (3.7%) | 4 (4.3%) | 6 (7.9%) | 6 (5.6%) | 4 (4.3%) | 26 (4.9%) |
| **BMI** |  |  |  |  |  |  |
| **Median [Q1, Q3]** | 26.1 [21.7, 31.2] | 31.6 [26.7, 38.1] | 29.2 [24.7, 33.3] | 29.3 [24.3, 34.5] | 28.1 [24.2, 33.5] | 28.6 [23.9, 33.9] |
| **Missing** | 2 (1.2%) | 0 (0%) | 0 (0%) | 1 (0.9%) | 2 (2.1%) | 5 (0.9%) |
| **Immunocompromised** | 49 (29.9%) | 31 (33.3%) | 15 (19.7%) | 40 (37.4%) | 33 (35.1%) | 168 (31.5%) |

### Antibiotic Stewardship Pattern in Treating Pneumonia of Different Etiologies

The median NAT score on the day of BAL sample collection was 2 for bacterial pneumonia, microbiology-negative pneumonia, and non-pneumonia control episodes, which corresponded to empirical coverage for HAP or VAP (Figure 3). While some of the non-pneumonia control episodes were adjudicated to be completely noninfectious (e.g. heart failure exacerbation), some of these episodes were adjudicated to co-exist with extrapulmonary infections (e.g. pyelonephritis). When we further stratified non-pneumonia control episodes based on the presence or absence of extra-pulmonary infection (n = 36 vs. n = 75), non-pneumonia control episodes with concurrent extra-pulmonary infection had a median NAT score of 2 on the day of BAL collection, while those without extra-pulmonary infection had a median score of 1 (*P* = 0.014, Mann-Whitney U test). For viral pneumonia and mixed bacterial/viral pneumonia, the median NAT score on the day of BAL collection was 1, which corresponds to targeted HAP or VAP coverage. Notably, an antibiotic de-escalation pattern was evident in all pneumonia etiologies except resistant bacterial pneumonia on day 1-2 following BAL collection. This pattern was also reflected by the difference in average NAT scores of each pneumonia etiology (Figure 4). For the viral pneumonia cases, complete cessation of antibiotics often occurred.

**Figure 3.**
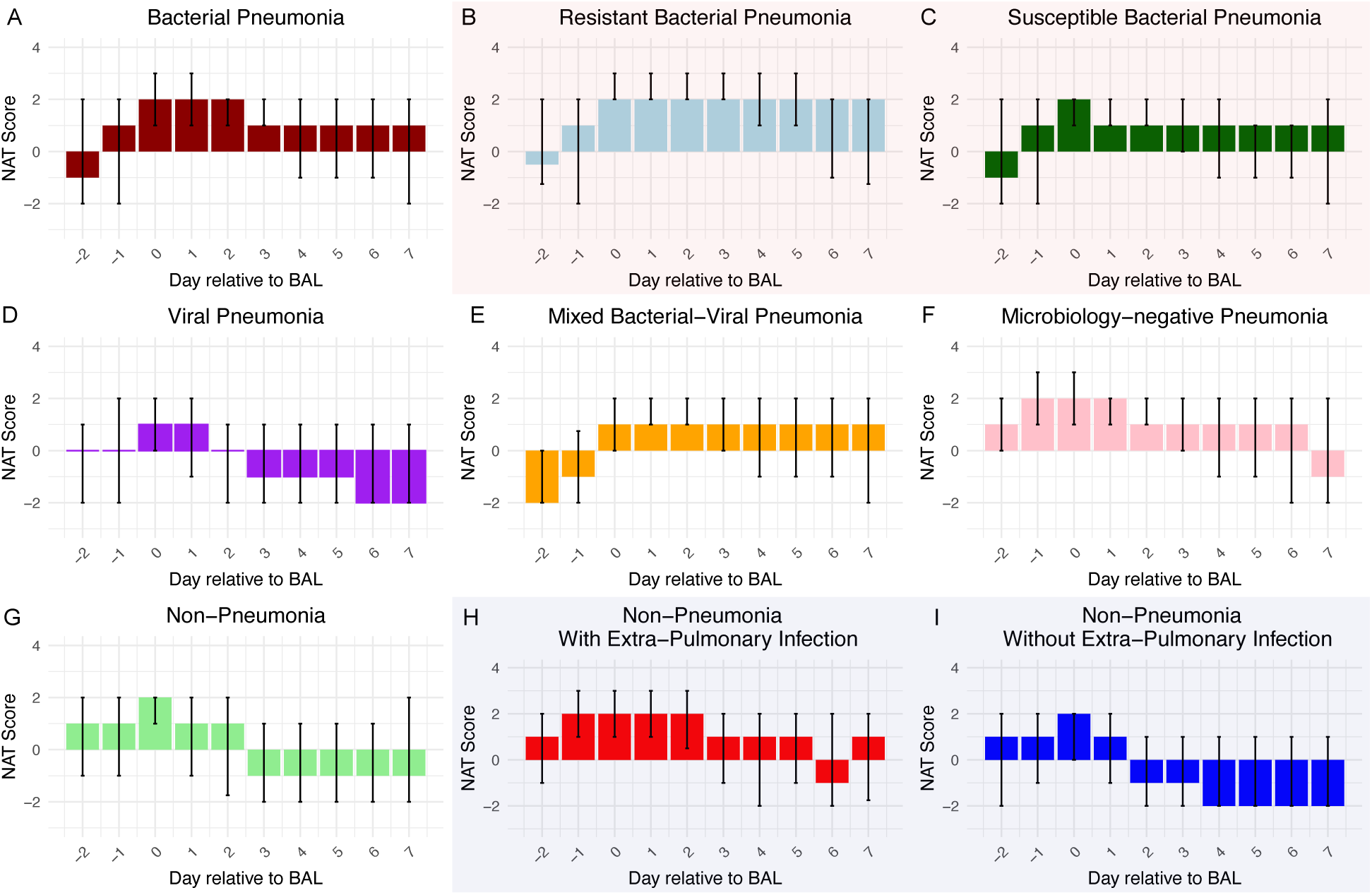
Variable patterns of antibiotic de-escalation, by category of pneumonia. Median NAT score per pneumonia episode day, with error bars representing IQR. If a patient died or was discharged before day 7, the days after death or discharge are not included in the plot. Plots B and C are subcategories of plot A, plots H and I are subcategories of plot G.

**Figure 4.**
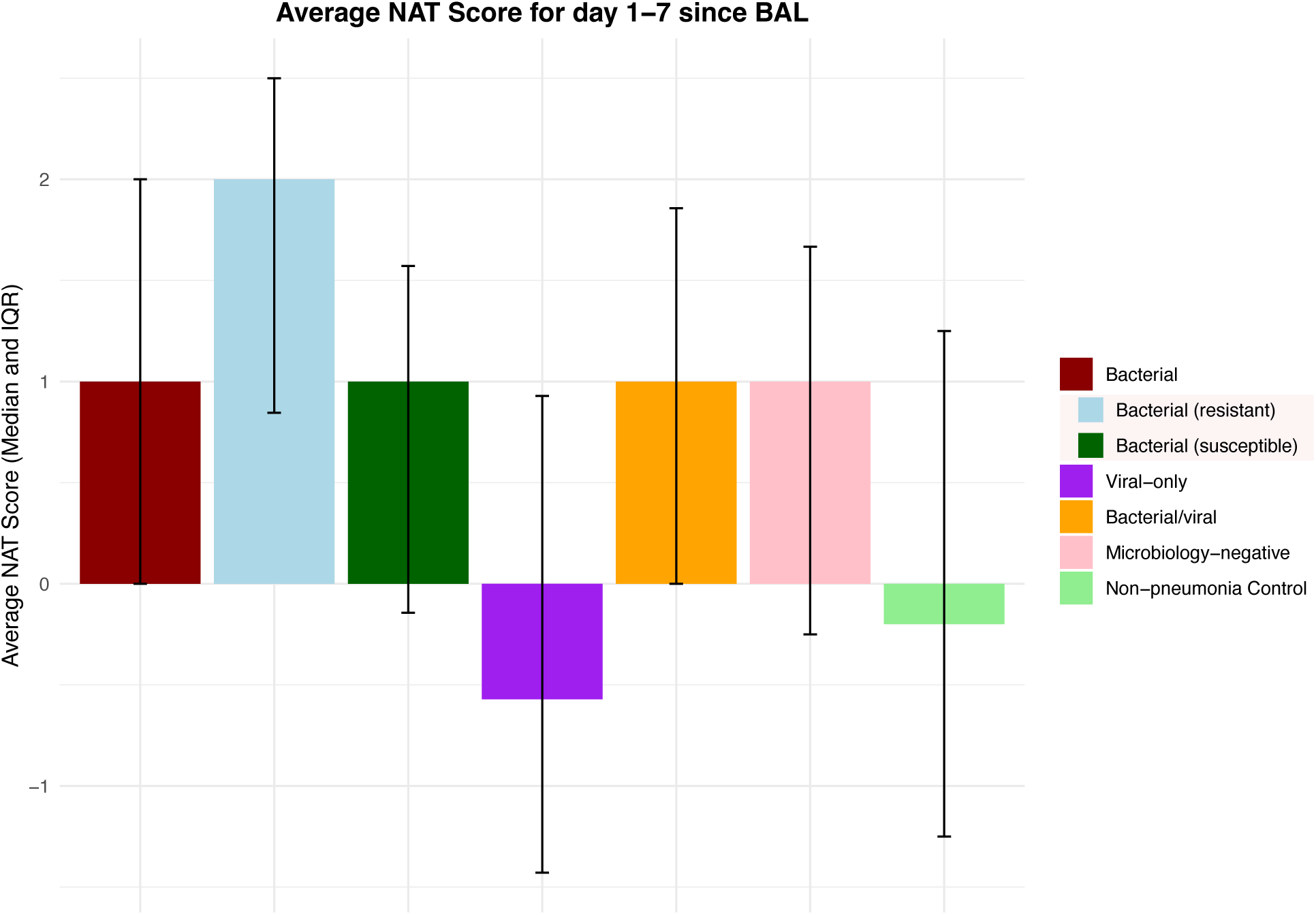
The average NAT score of days 1-7 relative to BAL collection shows the range of antibiotic de-escalation patterns across different episode etiologies. ‘Bacterial (resistant)’ and ‘bacterial (susceptible)’ are subcategories of ‘bacterial’ episodes.

To determine the association between pneumonia etiology, antibiotic stewardship patterns, and pneumonia outcome we further stratified bacterial, mixed bacterial/viral, and microbiology-negative pneumonia episodes based on the cure status on day 7 following BAL collection. Similarly, we stratified bacterial, viral, mixed bacterial/viral, microbiology-negative pneumonia episodes and non-pneumonia control episodes based on the presence or absence of concurrent extra-pulmonary infections. The cured pneumonia episodes of all etiologies demonstrated lower average NAT scores than uncured episodes (Figure 5). The episodes with concurrent extra-pulmonary infections demonstrate higher average scores, except for resistant bacterial pneumonia and mixed bacterial/viral pneumonia (Figure 6).

**Figure 5.**
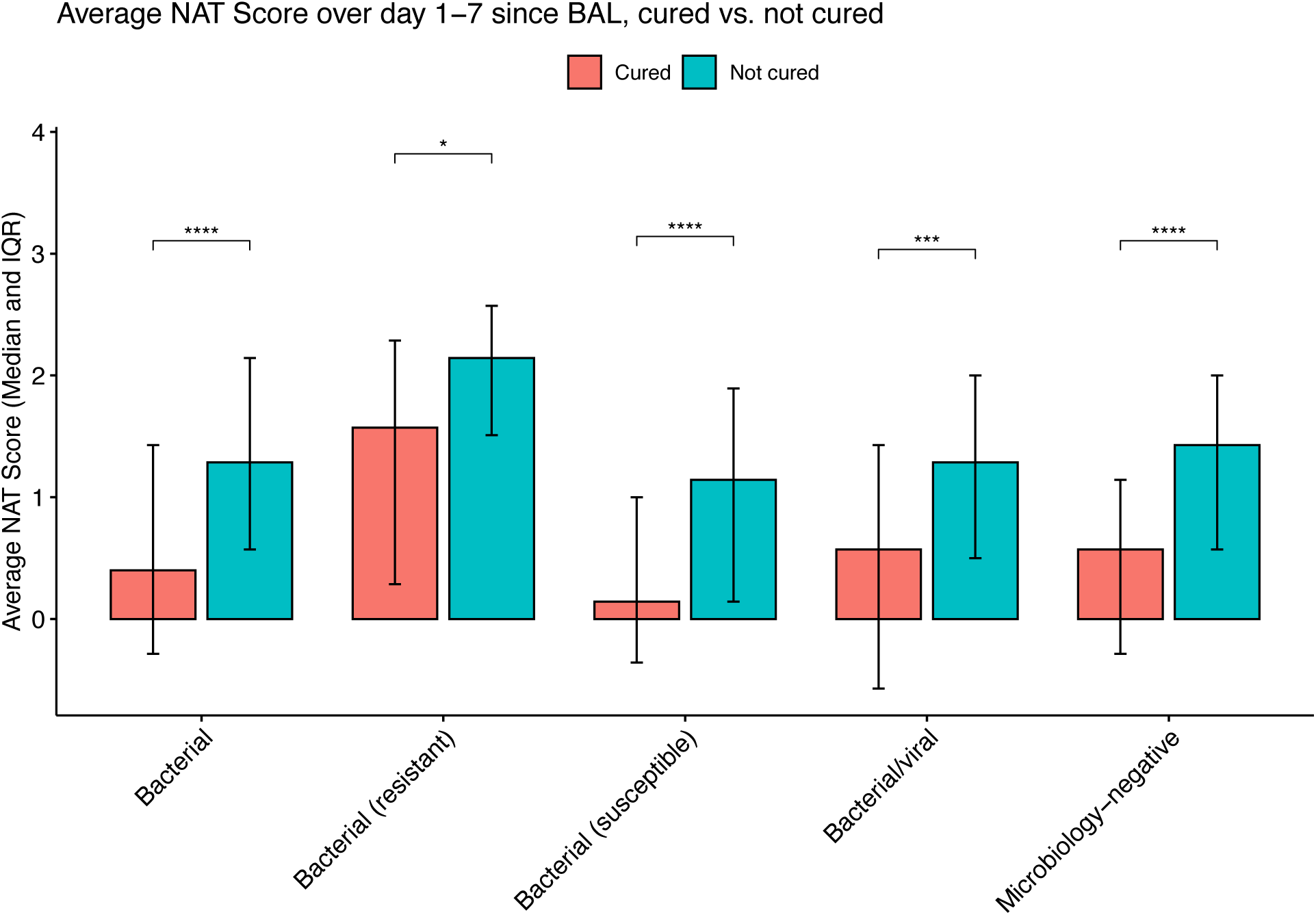
Average NAT score of days 1-7 relative to BAL collection, stratified by cure status on day 7. (*: P<0.05; **: P<0.01; ***: P<0.001; ****: P<0.0001)

**Figure 6.**
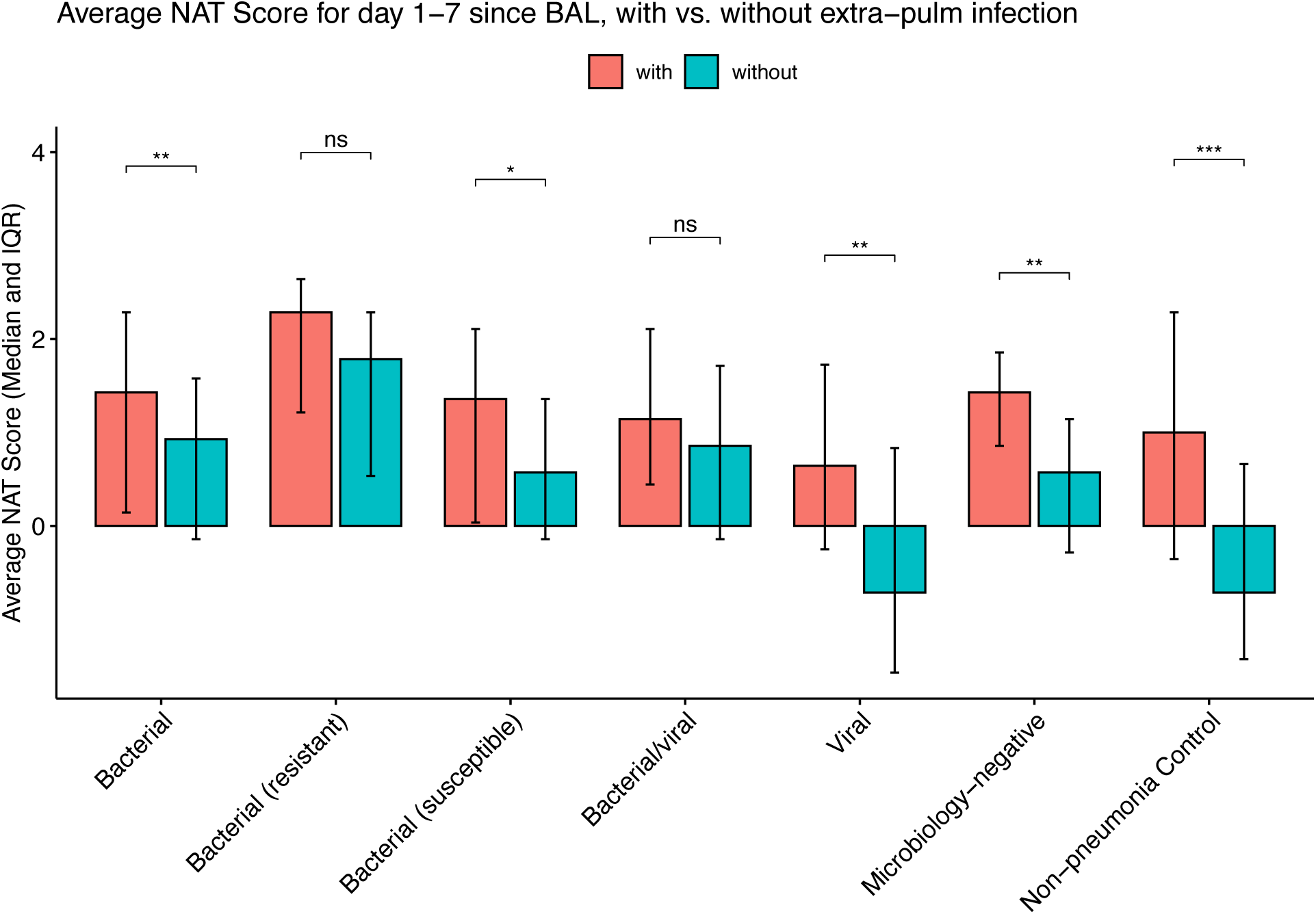
Average NAT score of days 1-7 relative to BAL collection, stratified by the presence or absence of extra-pulmonary infections. (ns: not statistically significant; *: P<0.05; **: P<0.01; ***: P<0.001; ****: P<0.0001)

### Clinical Outcomes

Unfavorable outcomes, duration of ICU stay, and duration of mechanical ventilation among patients with exactly one suspected pneumonia episode are reported in Table 3. All categories of pneumonia had comparable unfavorable outcome rates to microbiology-negative pneumonia (Bacterial: OR = 1, *P* = 1.00; Resistant bacterial: OR = 1.19, *P =* 0.71; Susceptible bacterial: OR = 0.94, *P =* 0.89; Viral: OR = 1.09, *P* = 0.78; Bacterial/viral: OR = 1.18, *P =* 0.65). Patients with bacterial pneumonia and microbiology-negative pneumonia had overall comparable durations of ICU stay and intubation, but patients with viral pneumonia and mixed bacterial/viral pneumonia had significantly longer durations of ICU stay and intubation than those with microbiology-negative pneumonia.

**Table 3.** Clinical outcomes in patients with exactly one pneumonia episode.

|  | Bacterial | Resistant | Susceptible | Viral | Bacterial/viral | Microbiology-negative | Non-Pneumonia Control | Overall |
| --- | --- | --- | --- | --- | --- | --- | --- | --- |
|  | (N=164) | (N=41) | (N=123) | (N=93) | (N=76) | (N=107) | (N=94) | (N=534) |
| Unfavorable outcome | 69 (42.1%) | 19 (46.3%) | 50 (40.7%) | 41 (44.1%) | 35 (46.1%) | 45 (42.1%) | 43 (45.7%) | 233 (43.6%) |
| Duration of ICU Stay (Median [Q1, Q3]) | 11.0 [7.00, 18.3] | 14.0 [8.00, 19.0] | 10.0 [7.00, 18.0] | 15.0 [10.0, 20.0] | 16.5 [11.0, 28.3] | 11.0 [6.00, 19.0] | 8.50 [5.00, 17.0] | 12.0 [7.00, 20.0] |
| Duration of Intubation (Median [Q1, Q3]) | 8.00 [5.00, 14.0] | 10.0 [5.00, 15.0] | 7.00 [5.00, 13.0] | 11.0 [7.00, 17.0] | 13.0 [6.00, 26.0] | 8.00 [4.00, 15.0] | 6.00 [3.00, 11.8] | 9.00 [5.00, 15.8] |

## Discussion

Our study describes an antibiotic prescription pattern where early antibiotic de-escalation is informed by PCR RDT and quantitative cultures performed on BAL samples. This study is one of the first to quantify the de-escalation of antibiotic therapy on consecutive ICU days with the NAT score. The NAT score distinguishes between broad-spectrum and narrow-spectrum antibiotics and is sensitive to the number of antibiotics given per day, providing more granular information compared with binary features such as antibiotic-free days. Results from our study suggest the feasibility and safety of antibiotic de-escalation in critically ill patients with microbiology-negative pneumonia and highlight BAL with PCR RDT and quantitative cultures can accelerate the time to de-escalation. We described an antibiotic management strategy from our institution where de-escalation occurred promptly after a BAL sample resulted in no resistant pathogens detected by multiplex PCR and quantitative cultures.[11] With early antibiotic de-escalation informed by BAL studies, we treated critically ill patients with microbiology-negative pneumonia similarly to or less aggressively than those with susceptible bacterial pneumonia and achieved comparable clinical outcomes. The presence of clinical improvement and extra-pulmonary infections is taken into consideration when implementing antibiotic de-escalation.

Due to their critical illness, clinical hesitancy to de-escalate antibiotics in ICU patients despite negative microbiology tests leads to inappropriately prolonged broad-spectrum antibiotic therapy.[23,24] The antibiotic prescription patterns described in this study suggest how BAL sampling encourages clinical practice largely consistent with the recommendations from the IDSA and enables early de-escalation of antibiotics following infectious workup without evidence of a resistant pathogen.[22] The recommendation on antibiotic stewardship in microbiology-negative pneumonia is a recent addition to the guidelines, as previous guidelines did not provide specific recommendations for this clinical scenario.[25] Despite this recent update to guidelines, many patients with microbiology-negative pneumonia do not receive antibiotic de-escalation in a timely fashion. In a retrospective cohort study, which included 164 hospitals across the United States, only one out of every seven patients with microbiology-negative pneumonia received antibiotic de-escalation by hospital day 4.[3]

Interestingly, the patients with viral-only pneumonia received the greatest antibiotic de-escalation (sometimes complete discontinuation) after BAL collection, which differs from the current published CAP guidelines that recommend continuing antibiotics in this scenario. The viral-only cases had the lowest average NAT score, and although the duration of mechanical ventilation and ICU stay were longer, still had comparable mortality to other categories of pneumonia. Our viral-only cases mostly comprised patients with SARS-CoV-2 pneumonia, which is known to have longer ventilation and ICU durations.[26][20]

Limitations to our study include its occurrence at a tertiary academic center with access to state-of-the-art rapid multiplex PCR, a robust BAL practice, including a significant NBBAL subset performed independently by respiratory therapists,[27] and an established comfort level with basing antibiotic management on test results.[13] This was an observational study, and thus reflect heterogeneity of practice by different physicians in terms of specific antibiotic choices and processes of care. Since clinical improvement was taken into consideration when implementing antibiotic de-escalation, it was possible that less aggressive antibiotic strategies were preferentially selected for patients with improving clinical trajectory.

## Conclusion

Here we quantify and describe day-by-day antibiotic de-escalation patterns from our institution in different etiologies of pneumonia episodes. Over a quarter of the time in viral pneumonia episodes, antibiotics were completely discontinued. Future prospective studies investigating the efficacy of a protocolized de-escalation strategy versus routine clinical care will be helpful to validate the feasibility and safety of more aggressive de-escalation.

## Human Ethics and Consent to Participate

This study was approved by the Northwestern University Institutional Review Board with study ID STU00204868. Study participants or their surrogates provided informed consent.

## Supporting information

STROBE checklist

Extended author list for NU SCRIPT Investigators

## Data availability

A significant portion of this data has been already made available through PhysioNet at https://physionet.org/content/script-carpediem-dataset/1.1.0/,[28] a future update will include new patients and updated data since the publication of the original dataset. Code is available at https://github.com/NUSCRIPT/mz_abx_deescalation_2024.

---

**Supplemental Table 1.**
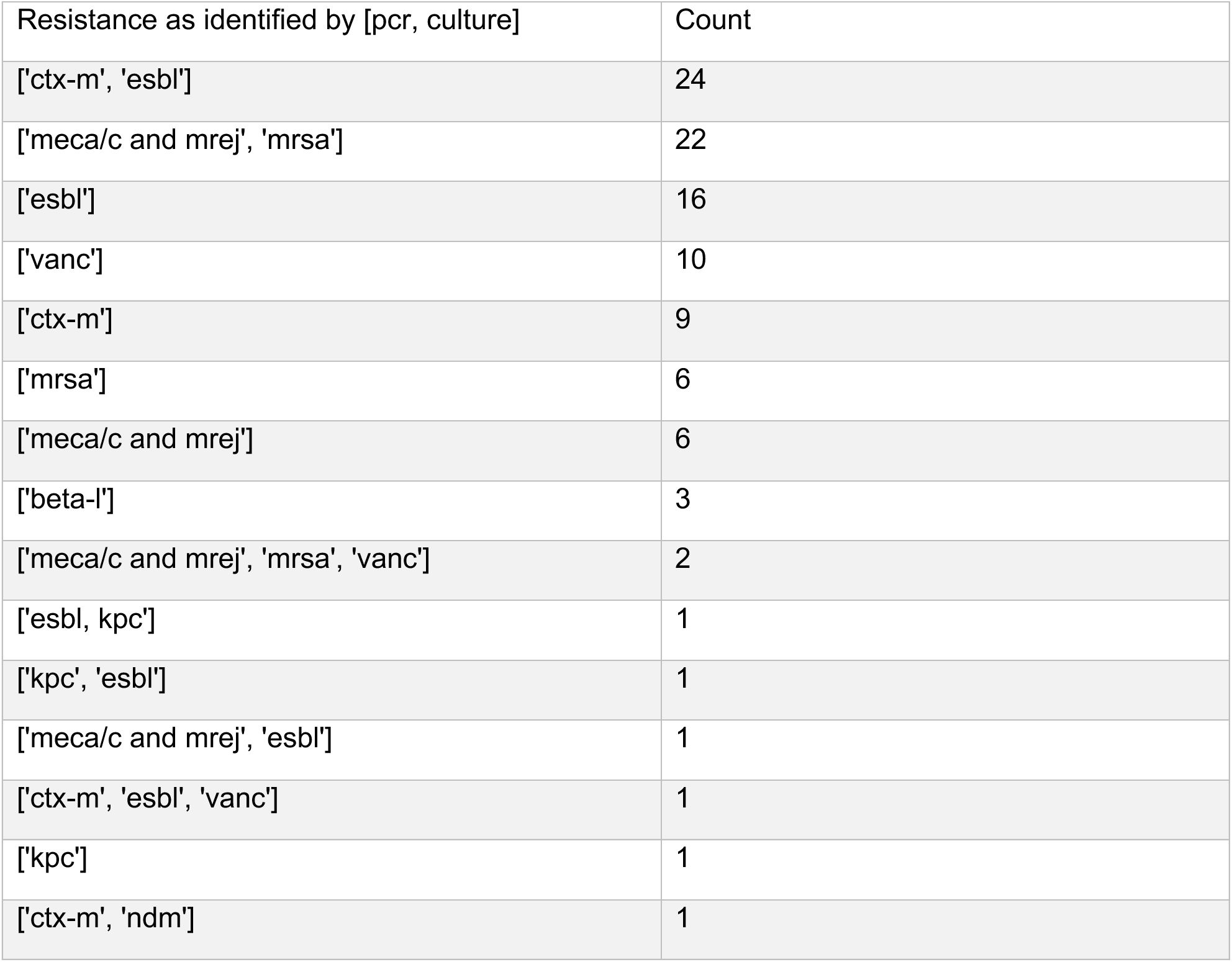
Summary of resistant pathogens as detected by either multiplex PCR mutation or in culture name and their frequencies. Pathogens were identified based on the presence of specific resistance genes or mechanisms. For example, [’meca/c and mrej’, ’mrsa’] indicates that a mecA/C and MREJ mutation were recovered on the Biofire panel, and the culture grew out MRSA. Abbreviations: CTX-M (cefotaximas-Munich enzyme causing EXBL phenotype), ESBL (Extended-spectrum β-lactamase), mecA/C (Methicillin resistance genes), MREJ (Methicillin resistance gene), MRSA (Methicillin-resistant Staphylococcus aureus), vanc (Vancomycin resistance), beta-L (Beta-lactamase), KPC (Klebsiella pneumoniae carbapenemase), and NDM (New Delhi metallo-β-lactamase).

**Supplemental Table 2.** Frequency of bacterial pathogens in the SCRIPT cohort.

| Pathogen | Frequency |
| --- | --- |
| ***Staphylococcus aureus*** | 121 |
| *Pseudomonas aeruginosa* | 89 |
| *Viridans streptococcus* | 49 |
| *Escherichia coli* | 47 |
| Yeast, not *Cryptococcus species* | 46 |
| *Staphylococcus coagulase negative* | 45 |
| *Enterococcus faecalis* | 41 |
| *Klebsiella pneumoniae* | 40 |
| *Corynebacterium species* | 36 |
| *Klebsiella aerogenes* | 21 |
| *Haemophilus influenzae* | 20 |
| *Enterobacter aerogenes* | 18 |
| *Enterobacter cloacae complex* | 18 |
| *Serratia marcescens* | 17 |
| *Enterobacter cloacae* | 16 |
| *Enterococcus faecium* | 16 |
| *Klebsiella oxytoca* | 11 |
| *Stenotrophomonas maltophilia* | 11 |
| *Streptococcus agalactiae* | 11 |
| *Citrobacter koseri* | 10 |
| *Lactobacillus species* | 8 |
| *Neisseria species* | 8 |
| *Streptococcus pneumoniae* | 7 |
| *Acinetobacter calcoaceticus-baumannii* complex | 6 |
| *Beta hemolytic streptococci, group f* | 6 |
| *Proteus spp.* | 6 |
| *Acinetobacter baumannii* | 5 |
| *Beta hemolytic streptococci, group c* | 5 |
| *Burkholderia cepacia* complex | 5 |
| *Stomatococcus species* | 5 |
| *Achromobacter xylosoxidans* | 4 |
| *Citrobacter freundii* | 4 |
| *Beta hemolytic streptococci, group g* | 3 |
| *Hafnia alvei* | 3 |
| *Streptococcus pseudopneumoniae* | 3 |
| *Candida albicans* | 2 |
| *Candida tropicalis* | 2 |
| *Chryseobacterium indologenes* | 2 |
| *Elizabethkingia meningoseptica* | 2 |
| *Enterococcus avium* | 2 |
| *Granulicatella adiacens* | 2 |
| *Legionella pneumophila* | 2 |
| *Moraxella catarrhalis* | 2 |
| *Achromobacter denitrificans* | 1 |
| *Achromobacter species* | 1 |
| *Acinetobacter ursingii* | 1 |
| *Actinomyces odontolyticus* | 1 |
| *Arcanobacterium haemolyticum* | 1 |
| *Beta-hemolytic streptococci, not group a, b, c, d, f, or g* | 1 |
| *Burkholderia gladioli* | 1 |
| *Candida auris* | 1 |
| *Candida glabrata* | 1 |
| *Chryseobacterium gleum* | 1 |
| *Enterococcus raffinosus* | 1 |
| *Enterococcus species* | 1 |
| *Granulicatella species* | 1 |
| *Morganella morganii* | 1 |
| *Neisseria meningitidis* | 1 |
| *Providencia stuartii* | 1 |
| *Pseudomonas fluorescens* | 1 |
| *Raoultella ornithinolytica* | 1 |
| *Streptococcus constellatus* | 1 |
| *Streptococcus mitis oralis* | 1 |
| *Streptococcus pyogenes* | 1 |

**Supplemental Table 3.** Breakdown of pneumonia categories and etiologies.

| **Etiology** | **Category** | **count** |
| --- | --- | --- |
| **Bacterial** | CAP | 48 |
|  | HAP | 83 |
|  | VAP | 157 |
| Bacterial/viral | CAP | 19 |
|  | HAP | 34 |
|  | VAP | 145 |
| Microbiology-negative | CAP | 34 |
|  | HAP | 72 |
|  | VAP | 45 |
| Viral | CAP | 49 |
|  | HAP | 68 |
|  | VAP | 59 |
| Non-pneumonia control | NPC | 114 |
| **Total** |  | 927 |

**Supplemental Table 4.** Frequency of Viral Pathogens in the SCRIPT Cohort.

| **Virus** | **Freq** |
| --- | --- |
| **SARS-Cov-2** | 303 |
| Influenza | 18 |
| Human<br>Rhinovirus/Enterovirus | 17 |
| Coronavirus (not SARS-Cov-2) | 13 |
| Parainfluenza | 9 |
| Adenovirus | 8 |
| Respiratory Syncytial Virus | 8 |
| Human Metapneumovirus | 5 |
| CMV | 3 |
| HSV | 3 |
| Herpes Zoster | 2 |

## Footnotes

* List of immunocompromised status/medications used by research team to flag patients as immunocompromised. Medical conditions: Acute leukemia, HIV, Immunoglobulin deficiency, Lymphoma, Multiple myeloma, Solid organ transplant, Stem cell transplant, Hematologic malignancy, Other malignancies/Non-malignancies. Medications: Azathioprine, Chronic corticosteroids (last month) > 5 mg/d, Chronic corticosteroids (last month) >= 20 mg/d, Cyclosporine, Cytoxan, Mycophenolate (MMF), Myelosuppressive chemotherapy, Rituximab, Tacrolimus, Other

